# PGxRAG: A Retrieval Augmented Generation supported Pharmacogenomics Assistant

**DOI:** 10.1101/2025.09.24.25336524

**Authors:** Dhanush Borishetty, Peter Banda, Nikhilesh Andhi, Aakash Desai, Bharath Ram Uppili, Naga Mithil Samudrala, Shravani Shriya Palanki, Dheeraj Reddy Bobbili, Gayatri Rangarajan Iyer

## Abstract

Pharmacogenomics enables personalized medicine by predicting individual drug responses based on genetic makeup, but complex guideline retrieval remains challenging for clinicians, particularly in resource-limited settings. While large language models (LLMs) show promise for numerous healthcare applications, their performance on domain-specific pharmacogenomics queries without expert knowledge integration remains limited. We evaluated whether Retrieval-Augmented Generation (RAG) enhancement improves LLM accuracy for pharmacogenomics applications compared to native model performance. We conducted comparative evaluation of four LLMs with and without RAG enhancement, constructing a knowledge base from PharmGKB (now ClinPGx), CPIC, Dutch Pharmacogenetics Working Group (KNMP), and FDA guidelines containing 2,617 embedded document chunks. We developed 225 multiple-choice questions representing patient and healthcare provider perspectives, then systematically evaluated hyperparameter combinations testing different temperatures, embedding dimensions, retrieval methods, and k-values with different sample sizes. RAG-enhanced models consistently outperformed native LLMs, with optimal configuration (GPT-4o) achieving 95.1% accuracy compared to 89.8% for the same native model. The RAG approach significantly enhances LLM performance in pharmacogenomics applications, providing a scalable solution for making complex pharmacogenomic guidelines accessible to healthcare providers while maintaining high clinical decision support accuracy.

## Introduction

Generative artificial intelligence (Gen AI) is a progressively trending phenomenon in several domains involving complex and layered decision making actions (Bohr and Memarzadeh, 2020). Healthcare is one such field where the utility of Gen AI is being increasingly explored (Raza et al., 2024). Machine learning algorithms can perform extensive dataset analysis and generate valuable insights (Chakraborty et al, 2024). Since its inception, machine learning models have evolved to perform unsupervised learning almost matching human acumen (Reddy, 2024).

Clinical decision-making involves weighing complex algorithms that process multilayered patient data from different time periods, multiple concurrent ailments, and consultations across various specialties. (Stafie et al, 2024). Supervised machine learning algorithm and validated Gen AI has potential to simplify this process to accelerate accurate and timely healthcare delivery (Clusmann et al, 2023).

Among the various types of Generative AI models, two categories are particularly well-suited for clinical decision-making projects: autoregressive models, which use large language models to predict outputs based on previous data, and flow-based models, which utilize change of variables formulas to model complex distributions. (Akhtar, 2024 and Reddy 2024). Autoregressive models leverage the transformer architecture, which has revolutionized natural language processing through its attention mechanism that dynamically weighs the importance of words and their contextual relationships while making sequential predictions (Yang and Lawson, 2023; Vaswani et al., 2017). This architecture enables the model to capture both local and long-range dependencies in text by allowing each token to attend to all previous tokens in the sequence, resulting in more coherent and contextually appropriate outputs. The self-attention mechanism, a key component of transformer-based autoregressive models, calculates relevance scores between all pairs of tokens, effectively learning which parts of the input are most important for predicting the next token in the sequence. This serves as an excellent model to retrieve and paraphrase guidelines and literature from medical knowledge base.

Documentations and administrative requirements consume around 25% of clinician’s workday and LLMs have the potential to fill this gap (Clusmann et al, 2023). Successful integration of Gen AI into healthcare includes data labeling, deployment, data corpus, technical standardization, facilitating novel human and AI interface, and governance (Clusmann et al, 2023 and Reddy 2024). Reinforcement learning from human feedback acts as safety guard rails to ensure safe and responsible operation.

Pharmacogenomics is a branch of personalized medicine that can predict the response of an individual to medications based on the underlying genetic makeup. Based on the genetic makeup, individuals can be classified as poor, intermediate, normal, rapid or ultrarapid metabolizers, which in turn categorizes them into the drug responder and non-responder categories (Oates and Lopez, 2018). However, the relationship between genetic makeup and its associated phenotype is far from straightforward (Heinen, 2010). Following large-scale population studies, such as genome-wide association studies (GWAS) and the 1000 Genomes Project, healthcare has experienced a revolution through the discovery of new genotype-phenotype correlations (Zheng-Bradle and Flicek, 2017). This has not only accelerated accurate diagnosis but has also paved the way for utilizing the allele frequencies for translational pharmacogenomics care improving the overall healthcare outcomes. Consortia and agencies like Clinical pharmacogenomics implementation consortium (CPIC), European Ubiquitous pharmacogenomics (UPGx), US Food and drug agency (FDA) have laid down several guidelines for informed and safe use of drugs based on the genotype of an individual (Kabbani et al, 2023). Pharmacogenomics Knowledge Base (PharmGKB) is an open resource that efficiently curates the guidelines and is a compendium for actionable pharmacogenomics (Whirl - Carrillo et al, 2021). The retrieval and cross reference of guidelines can often be overwhelming and time consuming, especially in resource restricted settings where pharmacogenomics is not part of standard clinical care.

The advent of large language models (LLMs) has ushered in a new era of potential for personalized medicine, offering unprecedented capabilities in processing and synthesizing vast amounts of biomedical information. Applying LLMs to pharmacogenomics dataset can translate the drug-gene pair knowledge to actual clinical care (Murugan et al, 2024). This can be further linked to the electronic health records including genome sequencing data of the patient to deliver a customized clinical management for every patient.

To address the challenges and explore the potential of Generative AI in personalized medicine, our study evaluates the performance of LLMs enhanced with pharmacogenomics context for a Retrieval-Augmented Generation (RAG) model. Our approach involves providing the LLM with context from authoritative sources namely CPIC, the Dutch Pharmacogenomics Working Group (DPWG), The French National Network of Pharmacogenetics (RNPGx) and FDA guidelines/label warnings.

By comparing native LLM and LLM enhanced with RAG, we aim to assess the impact of domain-specific knowledge integration on the accuracy and reliability of LLM-generated recommendations in personalized medicine. This evaluation is crucial in determining the readiness of LLMs for pharmacogenomics applications and identifying areas for improvement. Our study contributes to the growing body of research on the application of artificial intelligence in healthcare, with a specific focus on the potential of LLMs to enhance personalized treatment strategies and improve patient outcomes.

## Results

### Contexts

Approximately 319 JSON files of the drug and gene interactions of guidelines from the PharmCAT (source: PharmGKB) were downloaded, which were split into 363 chunks. Text was extracted from the downloaded DPWG (farmacogentica) guidelines PDF files (source: KNMP/DPWG) using AzureAI’s Document Intelligence and then were split into 693 chunks with each chunk representing a guideline. CSV files had 1561 chunks, where each row was considered a separate chunk, as it contained information on a particular type of the interaction. As shown in *Figure 1 (a)*, altogether, a total of 2,617 chunks with their metadata were stored in the Pinecone cloud as vectors, embedded using OpenAI’s “*text-embedding-3-large”* model with 3,072 dimensions.

**Figure 1.**
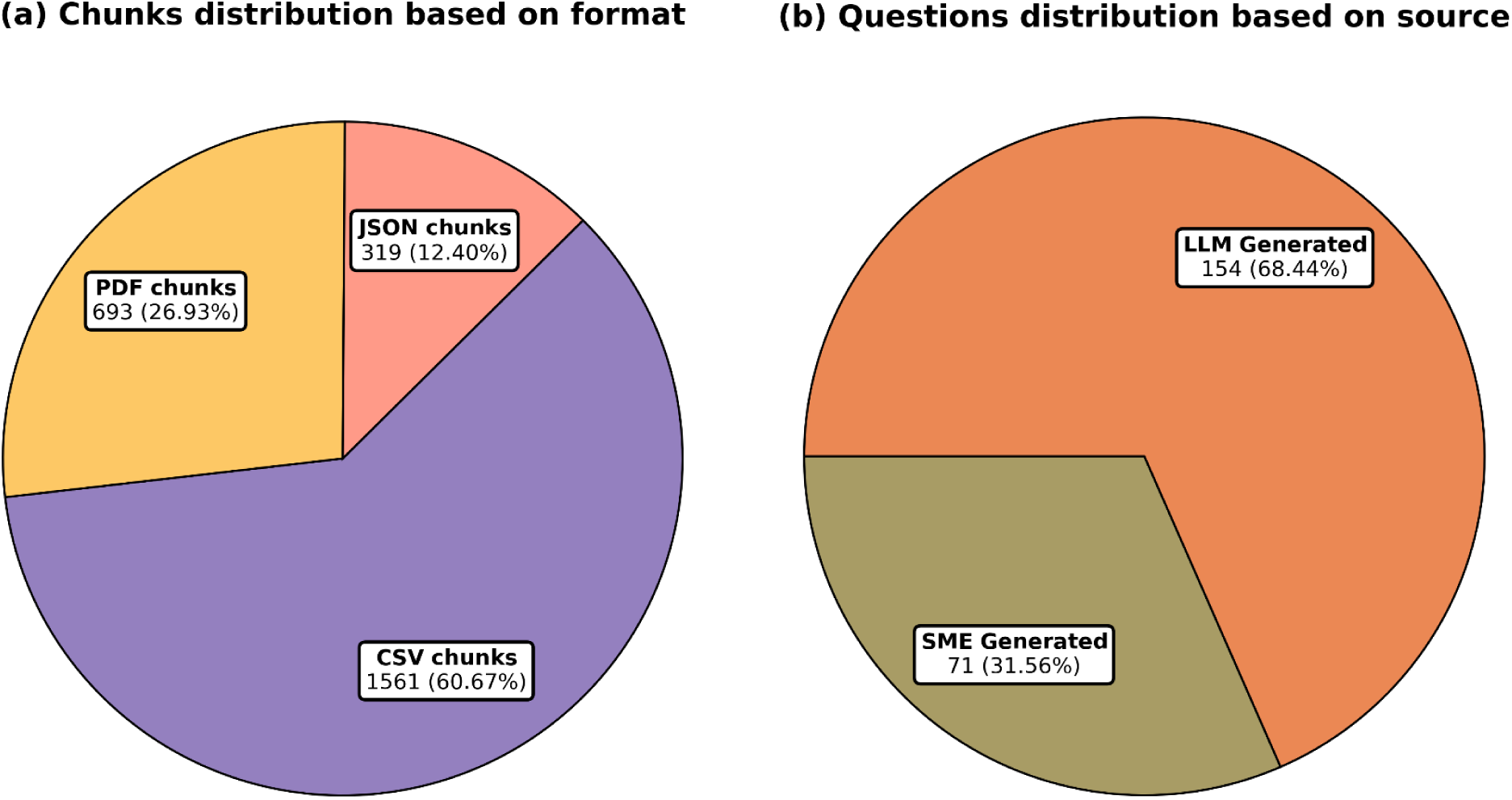
(a) Distribution of the chunks of guidelines based on the format (b) Distribution of the questions based on the source.

### Evaluation dataset

A total of 172 questions and answers were created by SMEs, of which 134 were converted into MCQ-type questions using GPT-4o, and 71 were selected by SMEs for further use. From the guidelines JSON files, 166 questions (18 + 148) were generated (one question per file), with 154 taken forward. As shown in *Figure 1 (b), a*ltogether a total of 225 (71 + 154) final set of questions were considered for the response generation and their evaluation by RAG (*Supplementary Table 1* has the list of questions with the ground truth/ expected or correct answer).

### Responses

Responses were generated (as shown in *Table 1*) by four LLM models - gpt-4o, gpt-4-turbo, claude-3.5-sonnet, llama3.1 (70b and 405b), which were assessed in different hyperparameter combinations for temperature (to limit creativity / follow probability distributions), embedding (for relevance and consistency), approaches like rerank (to enable reassessment and relevance), vector (for efficient retrieval in complex scenario) and hybrid (to improve accuracy) were applied, k value for (clustering information) to grasp the underlying patterns, error rate and accuracy. In total, 149 combinations were generated considering different sample sizes, as shown in *Supplementary Table 2*. From these results, it can be summarized that: (i) a temperature of 0 yielded better accuracy than 0.2; (ii) small embeddings (*text-embedding-3-small)* with 1,536 dimensions performed better than large embeddings with 3,072 dimensions (*text-embedding-3-large*) in OpenAI’s models; (iii) reranking improved response accuracy when combined with hybrid search; and (iv) lower k-values resulted in highers accuracy. As shown in *Figure 2*, among the different LLMs, Open AI’s GPT-4o (gpt-4o-2024-08-06) has the highest performance.

**Table 1:**
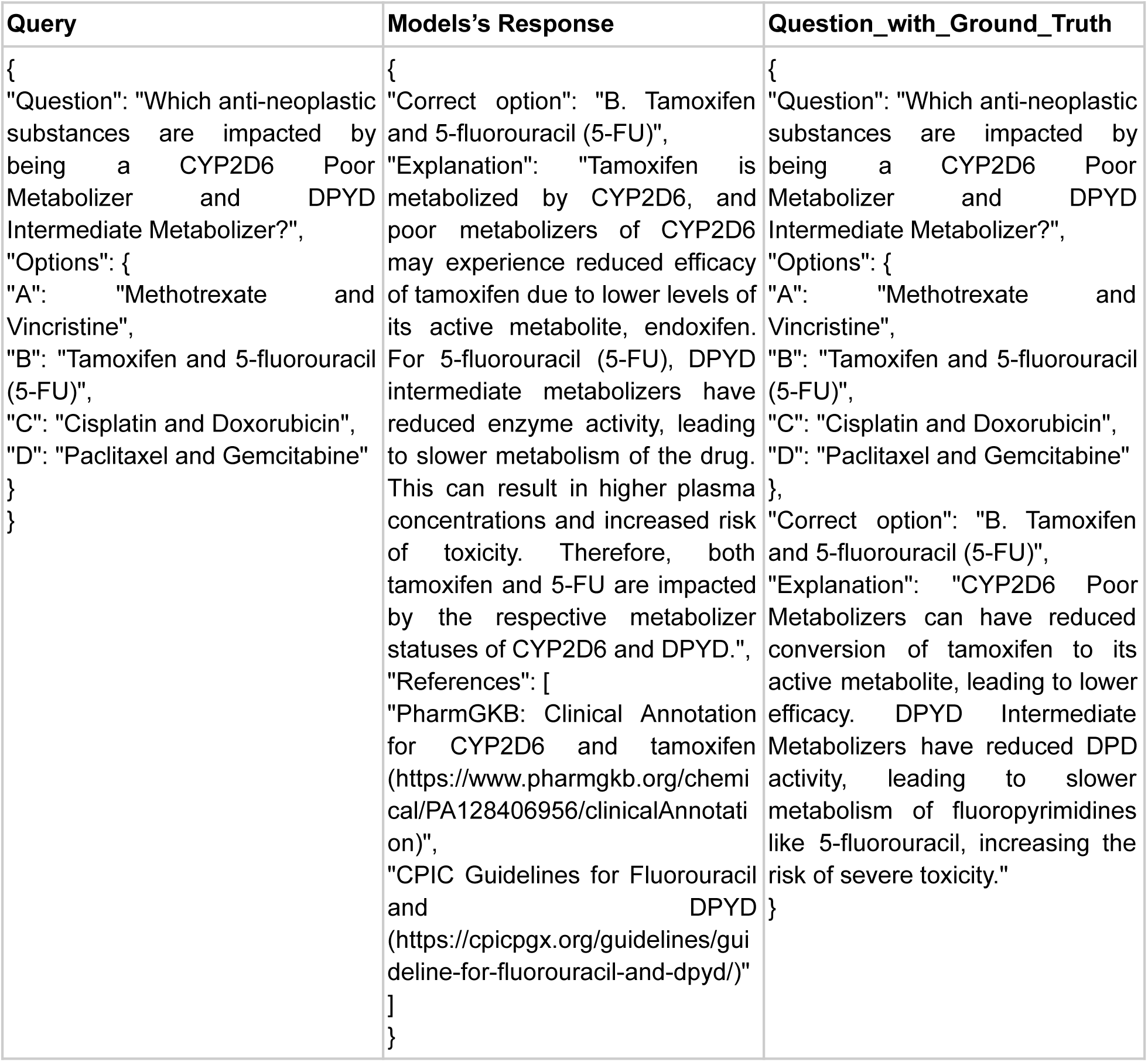
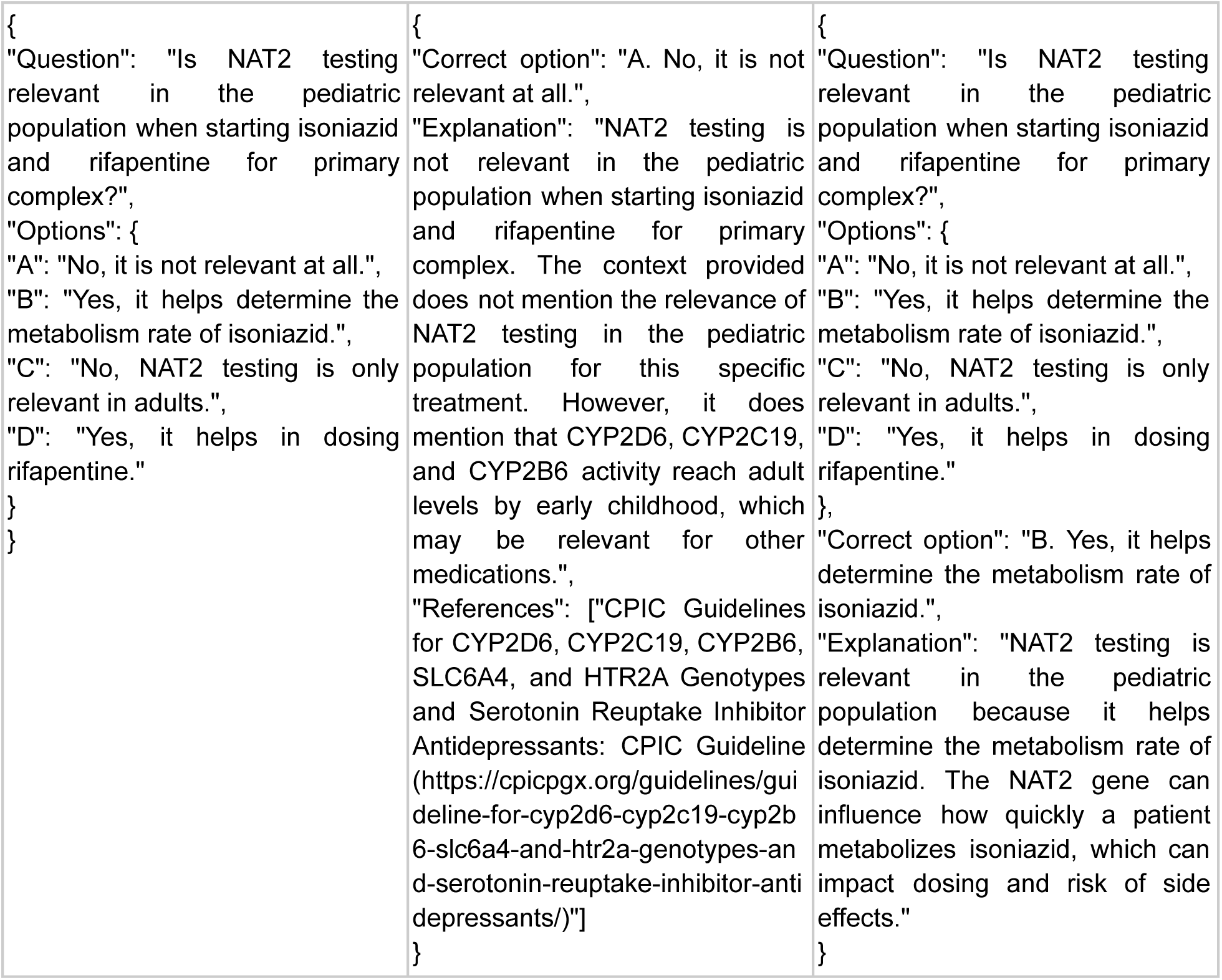
Question queried to a Model, its response, along with the Ground-Truth made (1st row of this table shows an example of model giving an expected correct answer, while the 2nd row is an example of model being wrong)

**Table 2:** Final hyperparametric combinations with corresponding Accuracy results.

| Model | Temperature | Embedding | Approach | k-value | Total | Correct | Accuracy |
| --- | --- | --- | --- | --- | --- | --- | --- |
| claude-3.5-sonnet | 0 | - | llm | - | 225 | 200 | 88.889 |
| claude-3.5-sonnet | 0 | small | rerank | 20 | 225 | 208 | 92.444 |
| claude-3.5-sonnet | 0 | small | rerank | 50 | 225 | 207 | 92.000 |
| gpt-4-turbo | 0 | - | llm | - | 225 | 182 | 80.889 |
| gpt-4-turbo | 0 | small | rerank | 20 | 225 | 209 | 92.889 |
| gpt-4-turbo | 0 | small | rerank | 50 | 225 | 211 | 93.778 |
| gpt-4o | 0 | - | llm | - | 225 | 202 | 89.778 |
| gpt-4o | 0 | small | rerank | 20 | 225 | 214 | 95.111 |
| gpt-4o | 0 | small | rerank | 50 | 225 | 213 | 94.667 |
| octoML-meta-llama-3.1-405b-instruct | 0 | - | llm | - | 225 | 181 | 80.444 |
| octoML-meta-llama-3.1-405b-instruct | 0 | small | rerank | 20 | 225 | 202 | 89.778 |
| octoML-meta-llama-3.1-405b-instruct | 0 | small | rerank | 50 | 225 | 201 | 89.333 |

**Figure 2.**
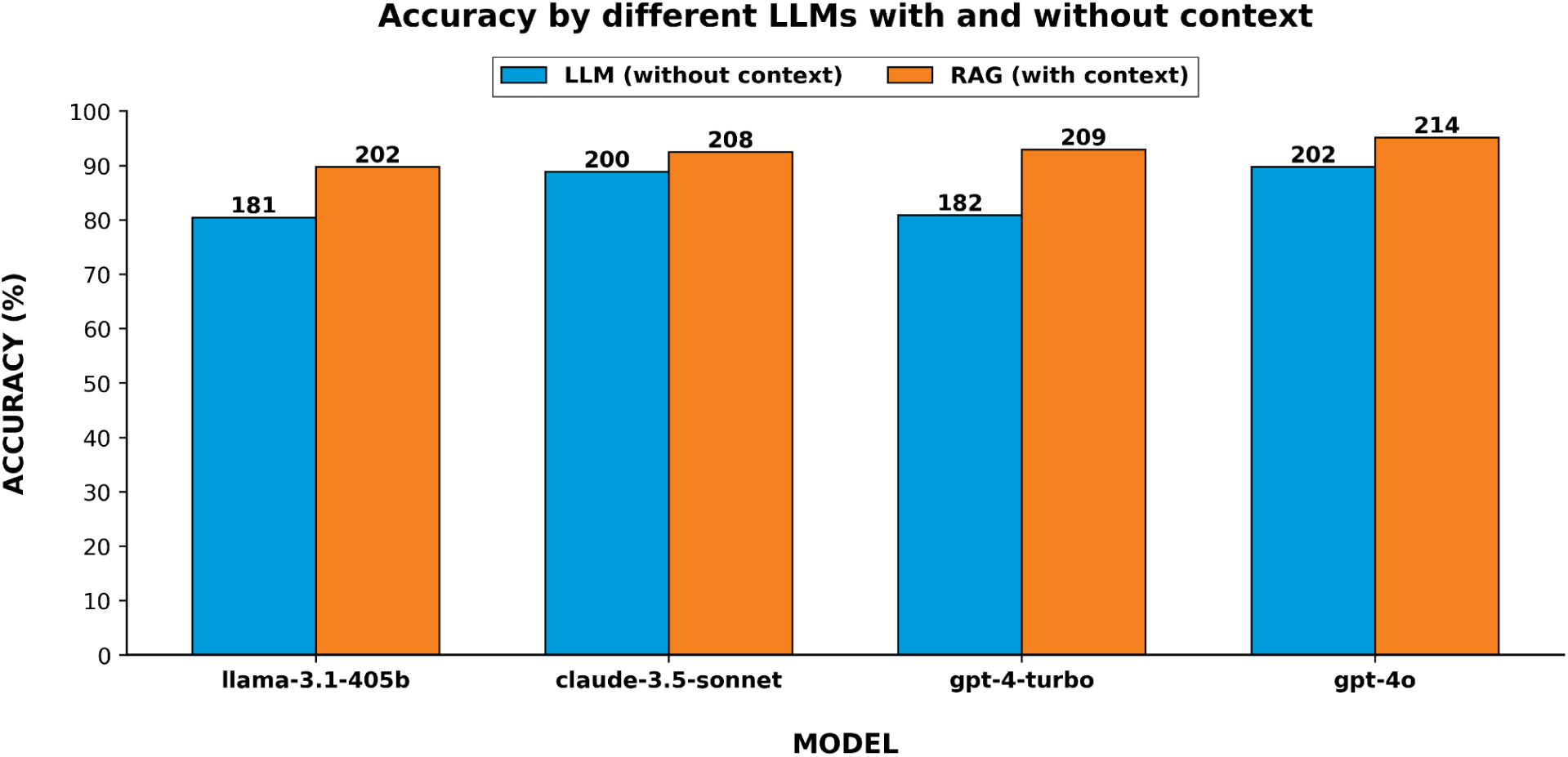
Comparison of Accuracy by LLM and best RAG hyperparameter combination for each of the models (the numbers on top of the bars indicate number of correct responses of the total 225 questions)

Accuracy of the models for correct responses were checked for both LLM and RAG approaches. The top four best performing models are demonstrated in *Figure 2*. RAG models (hyperparameter combinations) consistently performed better in each of the models. Model GPT-4o with RAG approach proved to be the best of the models with 95% accuracy. As shown in Table *2* and *Figure 2*.

## Discussion

Generative Artificial intelligence is increasingly being utilized in several domains including healthcare for prompt retrieval and summarization of data to facilitate faster and efficient delivery of services. Gen AI translation into healthcare can be adopted either at individual level which is quantifiable, addresses barriers, skill gaps and leads to seamless workflow integration or at organizational level where holistically multilevel functionalities and technical design is adopted (Reddy, 2024). Large language models like Generative Pre-training Transformer (GPT) are excellent context aware platforms for text summarization, if applied judiciously, can reduce about 25% of a clinician’s workday documentation, thus improving efficiency and preventing burnouts.

Pharmacogenomics is a field of personalized medicine which can help in predicting an individual’s response to medication based on their genetic makeup. There are different consortia and agencies that host the guidelines and information for clinically actionable pharmacogenomics, however, due to disparities in awareness, access, knowledge, cost and complex genotypic interpretations, pharmacogenomics has not been fully utilized in clinical care.

We have developed a LLM based pharmacogenomics assistant for easy referral and retrieval of drug-gene pair information from authoritative sources like PharmGKB, CPIC, FDA while accessing the electronic health records and genome sequencing data of the patient to interpret and summarize the pharmacogenomic guidelines based on patient genotypes/diplotype. The database is a contextual knowledge base with guidelines from CPIC, DPWG, FDA and review articles numerically embedded and LLM stored in a vectorbase.

Retrieval Augmented Generation (RAG) using vectorbase converted dataset and retrieved pertinent information based on the user queries using maximal marginal relevance (MMR) search. Questions were generated from patient and clinician perspectives for general pharmacogenomics knowledge, guidelines for specific drugs, concerns regarding adverse drug reactions and a few out of scope questions to assess the credibility of the AI assistant. Responses were assessed for accuracy, and relevancy. Using only LLM based and LLM with RAG approach, we assessed the performance and LLM with RAG approach performed better with 95.11% accuracy. Once the model was selected, we trained it on collected data and adjusted the parameters till it accurately predicted or generated appropriate outputs. This type of reinforcement learning from human feedback acts as safety guard rails to ensure safe and responsible operation to prevent AI hallucination. LLM with RAG approach models were further queried with fine tuning of different hyperparameters. PGx assistant with temperature of 0 small embeddings with 1536 dimensions, reranking approach when combined with the hybrid search and lower k-values yielded better accuracy compared to other models.

We present a novel LLM AI-PGx assistant, PGxRAG that is trained on authoritative pharmacogenomics guidelines that is comprehensive, elaborative, easy to query and allows retrieval of sources when prompted. The answers generated by the model are context aware as well as allows use of both generic and brand name of medications. As compared to other pharmacogenomics assistants, our model is comprehensive, easy to integrate into different specialties of medicine as it can summarize actionable pharmacogenomics of all available guidelines for both adults as well as pediatric population and not condition/drug specific. It captures information for all actionable pharmacogenomic variants with clinical evidence for a clinician to integrate in their practice. Diplotype is considered complicated while drawing pharmacogenomics inference and hence previous studies including Murugan et al utilized genotype for the querying, the developed model considers all genotype inputs like genotype, haplotype, diplotype as well as enzyme phenotype status for query and generates accurate outcome (Murugal et al, 2024). This feature is user-friendly wherein the end user need not go through complex databases and symbols to deduce the genotype/diplotype of the patient to interpret the pharmacogenomics report.

Current study involved integration of pharmacogenomics resources into the database for their easy referring and summarization.The objective and subjective comprehension of the model were analyzed sequentially and the model performs with 95% of accuracy across all the query types. We envision to add additional layers of drug-drug interaction in addition to the current drug-gene interaction as well as enrich the database with information from population genome studies to predict the outcomes from population allele frequencies which can educate the clinician on priority alleles/genes to assess for improving the drug responses and minimizing adverse drug events. We acknowledge PGxRAG is currently trained to choose the correct answer from multiple choice questions format similar to objective tests in medical school, to evaluate the model’s comprehension of the PGx queries. There are limitations in the study, as the assistant might not be integrated as a chatbot/plug-in with its current form to assist clinicians in comprehending raw genomics data, we would still need to use standard bioinformatics tools to call the star alleles and metaboliser status. However, it can be used to retrieve dosage instructions based on the guidelines. Further, it can be used to educate, train and evaluate clinical geneticists, genetic counselors, pharmacy students and clinicians. We intend to develop it into a chatbot as a subjective querying tool wherein individual questions will be answered using the RAG approach. We used Gen AI to generate questions in addition to the ones generated by subject matter experts which might skip a certain range of questions. To overcome this, we followed a strict reviewing process to cover wide topics of questions and to prevent data hallucination. Resources were additionally reviewed in order to verify the answers, and drug-gene pairs for which there is no pertinent evidence were also consulted in order to assess the model’s correctness. There are no global consensus and policies existing to regulate the use of GenAI in healthcare. We look forward to guidelines to assess our model. We propose integrating PGxRAG into electronic health records as well as a plugin for the bioinformatics pipelines of genome analysis to summarize the key findings and exercise caution with prescription medicine. As more real world data gets assessed in PGxRAG, the robustness and accuracy of the model will improve.

### Conclusion

We have developed a PGxRAG, a pharmacogenomics assistant to query widely used databases. The input for the query could be subject related, general knowledge, concerns or dosage related in alternable forms of genotype, diplotype, phenotype or allelic information and the assistant will recognize the underlying genetic makeup and retrieve associated guidelines and recommendations. The LLM-RAG supported model has 95% accuracy in performance compared to other models and is comprehensive for all drugs having actionable information in PharmGKB, CPIC, RNPGx, FDA guidelines. The second version of the model will be inclusive of drug-drug interaction in addition to its current drug-gene information and will have a chatbot model to assist clinicians at the interface of electronic medical records, genome data for subjective querying and retrieval of guidelines.

## Methods

### Study Design

We conducted a comparative evaluation of Large Language Models (LLMs) in the domain of personalized medicine, focusing specifically on pharmacogenomics by probing the models with multiple choice questions and assessment of the responses. The study aimed to assess the performance of native LLMs versus Retrieval-Augmented Generation (RAG)-enhanced LLMs that incorporated external pharmacogenomic resources. The primary objective was to determine whether the integration of RAG could improve the accuracy, relevance, and clinical utility of LLM-generated responses to pharmacogenomic queries. The workflow of the methodology can be referred to at *Figure 3*.

**Figure 3.**
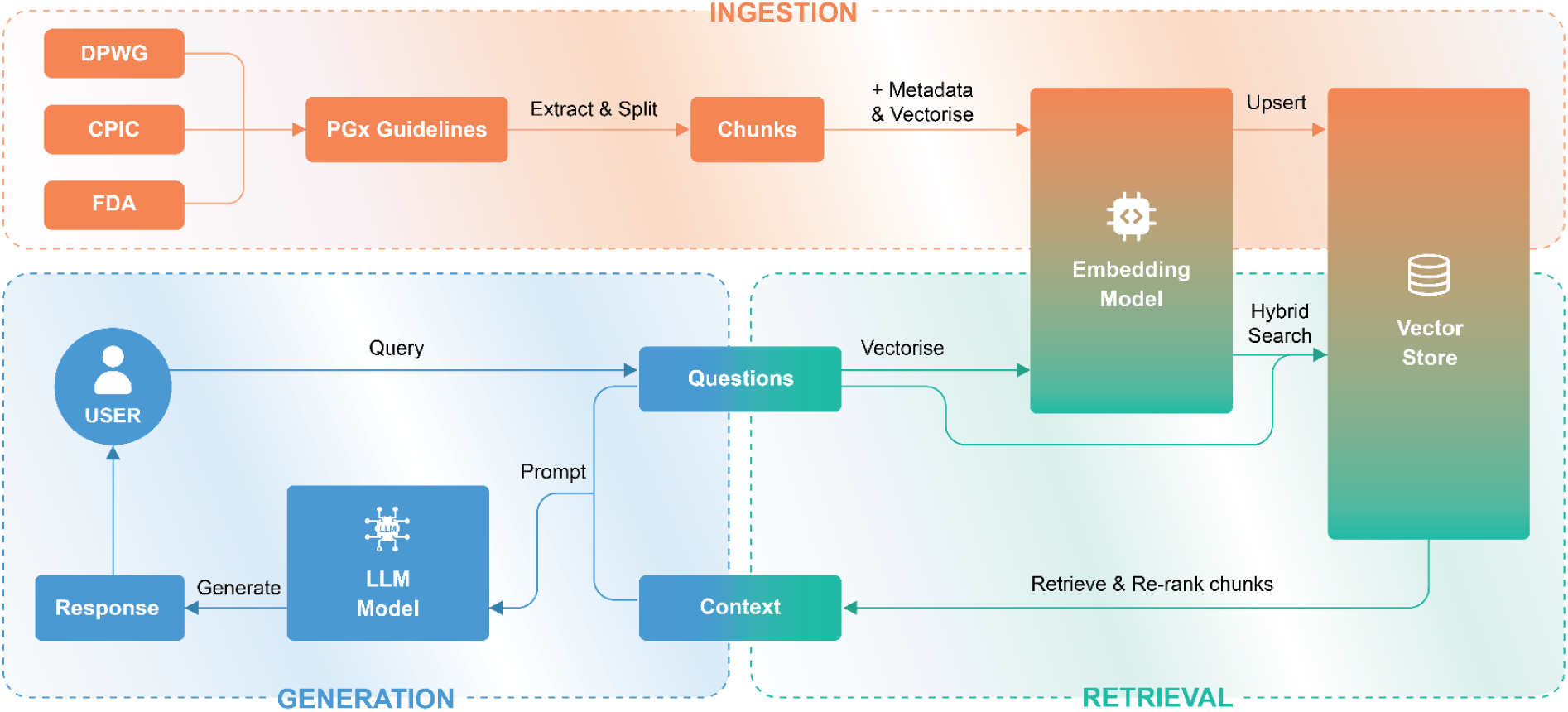
Schematic representation of RAG flow (i) INGESTION: Creating a vectorstore with embedded document chunks along with their metadata (ii) RETRIEVAL: Retrieving of re-ranked relevant chunks for the question queried against the vectorstore using hybrid search (iii) GENERATION: Using the question and retrieved relevant chunks, generating the response by the LLMs (iv) Accuracy evaluation by comparing the generated responses with the ground truth

### Evaluation Dataset Generation

A comprehensive set of multiple-choice questions (MCQs) related to pharmacogenomics was developed to simulate real-world clinical scenarios and inquiries from both patients and healthcare providers. These questions were prepared by Wellytics’ subject matter experts (SMEs) in pharmacogenomics, NA and GI.

The questions were categorized into two main perspectives: patient perspective and healthcare provider perspective. Each perspective was further subdivided into five categories:

1. Concerns: Reflecting common concerns from patients or providers regarding pharmacogenomics.
2. Pharmacogenomic Guideline-Specific: Focusing on the interpretation and application of pharmacogenomic guidelines.
3. Drug-Gene and Drug Interaction: Examining knowledge of drug-gene interactions and potential drug-drug interactions.
4. Educational/Generalized Pharmacogenomics: Testing general understanding of pharmacogenomic principles.
5. Out-of-Scope Queries: Assessing the models’ handling of irrelevant or unexpected queries.

The questions incorporated various clinical and demographic factors, including age, gender, ethnicity, side effects, past medication history, past medical history, family history, genes, gene variants (genotypes, alleles, haplotypes, diplotypes, rsID), metabolic phenotypes, applicable pharmacogenomic guidelines, types of adverse drug reactions, drug safety concerns, and alternative medications for unfavorable genotypes. Both generic and brand-name medications were included to assess the models’ recognition of different nomenclatures. At several instances, resources were also queried to validate the responses as well as drug-gene pairs where there is no actionable evidence were also queried to analyze the accuracy of the model.

To enhance the diversity and robustness of the question set while preserving data anonymity, Generative Adversarial Networks (GANs) were employed to generate additional questions. OpenAI’s GPT-4 was also utilized to generate MCQs with four options, including the correct answer. All generated questions underwent rigorous verification by the SMEs for contextual relevance, option relevance, and accuracy. Only questions that met these criteria were included in the final dataset which were 225.

### Construction of the Contextual Knowledge Base

We constructed a contextual knowledge base using authoritative pharmacogenomic resources, including the Pharmacogenomics Knowledgebase (PharmGKB), Clinical Pharmacogenetics Implementation Consortium (CPIC) guidelines, the Dutch Pharmacogenetics Working Group (DPWG) guidelines, and the U.S. Food and Drug Administration (FDA) pharmacogenomic associations.

Data from these sources were collected and processed into appropriate formats. Files were chunked differently depending on their format: CSV files were split by rows, PDF files were segmented based on headings and subheadings, and JSON files were chunked based on size limitations, ensuring each chunk contained information about a single interaction or guideline when possible. Each chunk was annotated with metadata, including source, drug names, gene names, and relevant clinical annotations, to facilitate accurate retrieval.

### Embeddings and Vector Store Creation

Each chunk of data was vectorized to create embeddings using OpenAI’s embedding models. We utilized the “text-embedding-ada-002” model with 1,536 dimensions and a larger embedding model with 3,072 dimensions. These embeddings were stored in Pinecone, a vector database designed for efficient similarity searches. The use of embeddings allowed for semantic similarity searches between the queries and the knowledge base.

### Retrieval-Augmented Generation Framework

We evaluated the performance of several LLMs in conjunction with the RAG framework. The LLMs tested included OpenAI GPT-4-Turbo, OpenAI GPT-4o, Anthropic Claude-3.5-Sonnet, and Meta AI’s Llama 3.1 with 70 billion and 405 billion parameters.

Different retrieval strategies were employed, including vector search using cosine similarity, hybrid search combining vector search with BM25 keyword search, and hybrid search incorporating Cohere’s reranking. We experimented with following hyperparameter combinations:

- Temperatures: Controlling randomness in model responses (temperature = 0, 0.2).
- Embedding Models: Using both the small (1536 dimensions) and large (3072 dimensions) embeddings .
- Retrieval Methods: Testing each strategy individually and in combination. Vector search by cosine similarity, Hybrid search using Vector search combined with BM25 keyword search, and Hybrid search combined with Cohere’s reranking
- k-values: Number of top documents retrieved (k = 20, 50, 100, 200, 250).

All combinations were systematically evaluated to determine the optimal configuration.

### Response Generation and Evaluation

For each question, relevant context was retrieved from the knowledge base using the specified retrieval strategy and hyperparameters. Several times, resources were additionally reviewed in order to verify the answers, and drug-gene pairs for which there is no pertinent evidence were also consulted in order to assess the model’s correctness. The LLM generated a response by selecting one of the four MCQ options, utilizing the retrieved context.The response of all different combination of parameters was taken where the RAG gave the correct option as response, these were cross-checked with the correct option which was initially generated, and each combination of parameter was given an accuracy scores (where number of correct option responses matched with the LLM generated correct option).

To also verify the usefulness of the RAG, we compared with just the LLMs responses accuracy without providing them with the context (native LLMs) on the same set of questions. Whichever set of RAG hyper parameter combinations resulted in higher accuracy was taken forward.

The responses were evaluated based on accuracy. Accuracy was determined by comparing the model-selected option with the correct answer verified by the SMEs.

## Data availability

The PGx Guideline files used and the final list of questions evaluated on for the current study are available in the huggingface repository Wellytics/PGxRAG and can be accessed via this link https://huggingface.co/datasets/Wellytics/PGxRAG.

## Code availability

The code for this study is available as a python notebook on huggingface repository Wellytics/PGxRAG and can be accessed via this link https://huggingface.co/datasets/Wellytics/PGxRAG.

## Acknowledgments

We acknowledge Wellytics Technologies Pvt. Ltd. for supporting computational infrastructure to carry out the work. We thank Ravikiran Gujarati for graphically designing the study flowchart.

## Author contributions

DB - Development, evaluation of the LLM-RAG model, data analysis, co-wrote the first draft of the manuscript. PB - Guidance in the development, review of Manuscript. NA - Subject matter expert to generate questionnaire. AD - Review of manuscript. BRU - Review of manuscript and data analysis. NMS - Review of manuscript. SSP - Review of manuscript. DRB - Conceptualisation, supervision and review of manuscript. GRI - Subject matter expert to generate questionnaire, co-conceptualisation, co-wrote the first draft of the manuscript and data analysis. All authors have read and approved the final version of the submitted manuscript.

